# Comparison of post-COVID depression and major depressive disorder

**DOI:** 10.1101/2021.03.26.21254425

**Authors:** Roy H. Perlis, Mauricio Santillana, Katherine Ognyanova, Jon Green, James Druckman, David Lazer, Matthew A. Baum

## Abstract

**Background:** During the COVID-19 pandemic rates of depressive symptoms are markedly elevated, particularly among survivors of infection. Understanding whether such symptoms are distinct among those with prior SARS-CoV-2 infection, or simply a nonspecific reflection of elevated stress, could help target interventions.

**Method:** We analyzed data from multiple waves of a 50-state survey that included questions about COVID-19 infection as well as the Patient Health Questionnaire examining depressive and anxious symptoms. We utilized multiple logistic regression to examine whether sociodemographic features associated with depression liability differed for those with or without prior COVID-19, and then whether depressive symptoms differed among those with or without prior COVID-19.

**Results:** Among 91,791 respondents, in regression models, age, gender, race, education, and income all exhibited an interaction with prior COVID-19 in risk for moderate or greater depressive symptoms (p<0.0001 in all cases), indicating differential risk in the two subgroups. Among those with such symptoms, levels of motoric symptoms and suicidality were significantly greater among those with prior COVID-19 illness. Depression risk increased with greater interval following acute infection.

**Discussion:** Our results suggest that major depressive symptoms observed among individuals with prior COVID-19 illness may not reflect typical depressive episodes, and merit more focused neurobiological investigation.

## Introduction

Rates of major depressive symptoms are elevated following acute infection with SARS-CoV-2(1)(2)(3). A key question is whether such symptoms represent a general consequence of stress associated with acute illness, or whether they may reflect more specific sequelae related to COVID-19 pathophysiology itself. To examine this possibility, we compared features of major depression in individuals with or without prior COVID-19 illness.

## Method

### Study design and cohort description

We conducted 12 waves of an internet non-probability Qualtrics survey using a multi-panel commercial vendor (PureSpectrum) approximately every month between May 2020 and February 2021 in individuals 18 and older. The study was approved by the Institutional Review Board of Harvard University. All participants provided informed consent. We followed AAPOR reporting guidelines, as described at https://www.aapor.org/Standards-Ethics/AAPOR-Code-of-Ethics/Survey-Disclosure-Checklist.aspx.

The survey included sociodemographic questions to identify gender, income, age, education, urbanicity, and self-identification of race/ethnicity, the last using 5 US Census categories, to ensure representativeness of the US population. It asked respondents if and when they had received a clinician diagnosis of COVID-19 illness or a positive SARS-CoV-2 result. Participants also completed the PHQ-9 as a measure of depressive symptoms(4), with 10 or greater defined as at least moderate depression.

### Analysis

We applied multiple logistic regression with moderate or greater depression as the dependent variable and sociodemographic features as independent variables, testing an interaction term with prior COVID-19 for each feature, using *glm* (R 3.6). We compared mean values for each depressive and anxious symptom between those with or without prior COVID-19, using Student’s t-test. Finally, we examined prevalence of depression by months elapsed since COVID-19.

## Results

Among 91,791 individuals who completed the PHQ-9, 61,472 (67.0%) were female, 9,667 (10.5%) Black, 6,686 (7.3%) Hispanic, and 5,306 (5.8%) Asian; 36,818 (40.1%) completed some college; 22,216 (24.2%) were in an urban location, and 16,158 (17.6%) rural. Mean age was 42.34 (SD 16.36), mean income $65,200 (SD $70,600) and 5,945 (6.5%) reported prior COVID-19 clinician diagnosis or test result while 28,617 (31.2%) reported moderate or greater depression.

In regression models, age, gender, race, education, and income all exhibited an interaction with prior COVID-19 in risk for depression (p<0.0001 in all cases; Figure 1). Comparing individual symptoms among those respondents with or without prior COVID-19, greatest differences were observed for suicidality (mean 2.50 (SD 1.06) versus 1.99 (SD 1.09); T=-24.83, p<0.0001) and motor symptoms (mean 2.58 (SD 1.01) vs 2.11 (SD 1.06); t = −23.74, p<0.0001) (Figure 2). Risk for depression increased with greater duration after acute illness (for each additional month, crude OR=1.07, 95% CI 1.1.05-1.09; adjusted for survey month, state, and sociodemographic features, aOR=1.09, 95% CI 1.07-1.11).

**Figure 1.**
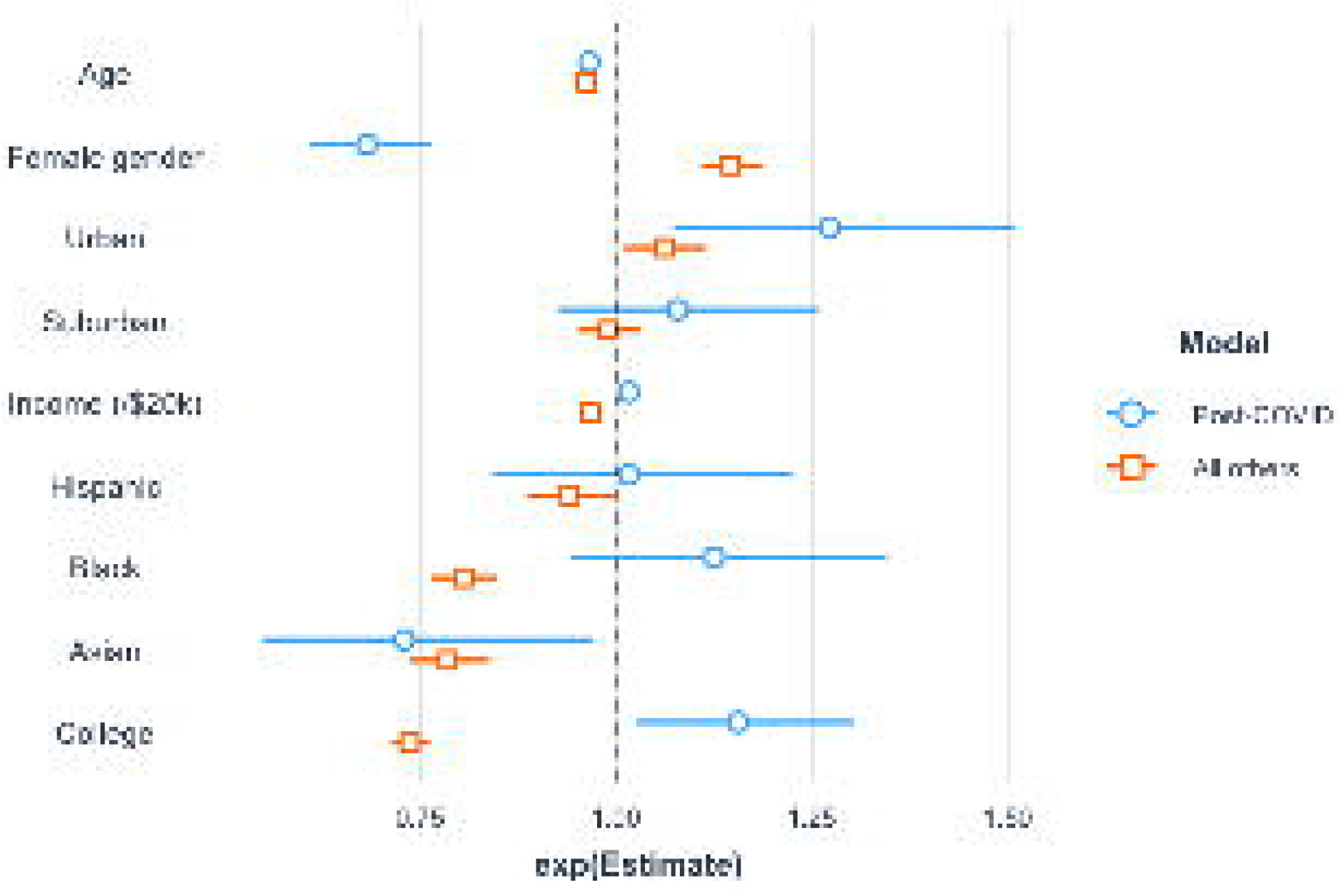
Logistic regression model for association of sociodemographic features with moderate or greater depressive symptoms, among those with or without prior COVID-19 illness

**Figure 2.**
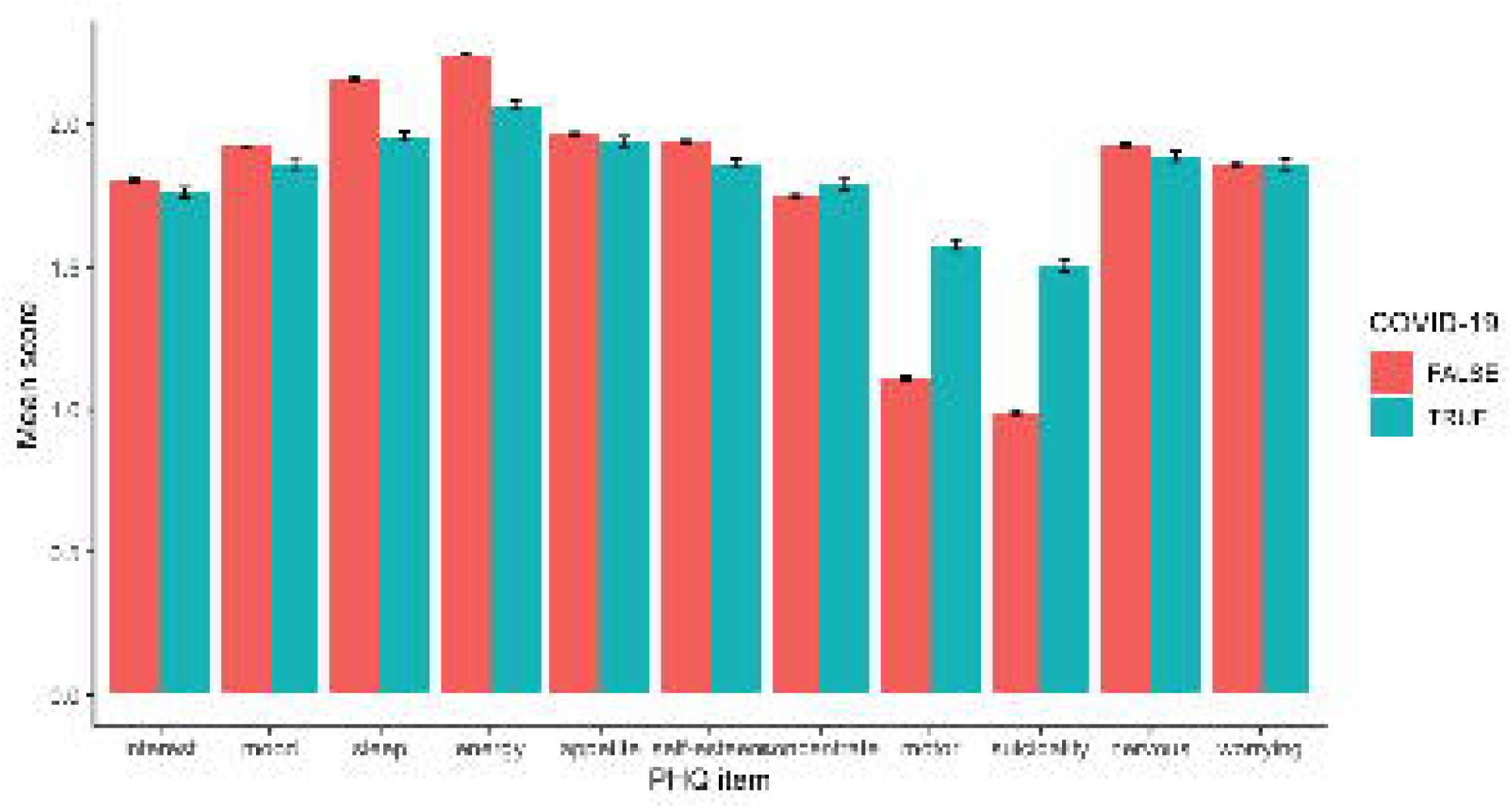
Individual PHQ-9 and PHQ-4 symptoms among individuals with current moderate or greater depressive symptoms, with and without prior COVID-19 infection

## Discussion

Magnitude of depressive symptoms and sociodemographic correlates differ between individuals with and without prior COVID-19 illness. These differences in phenomenology and risk factors both indirectly suggest that apparent major depressive episodes following COVID-19 illness may be distinct from those typically observed in adults. Furthermore, risk for depression increases with greater post-acute interval, rather than the gradual reduction anticipated if depression is simply a consequence of acute increase in illness-associated stressors.

The difference in symptom profile is notable in light of the markedly elevated rates of delirium observed in COVID-19 patients(5,6), given that delirium is also often associated with motoric symptoms as well as cognitive symptoms. On the other hand, the lesser differences in concentration and fatigue suggest that post-COVID-19 depression is not entirely explainable by other post-acute systemic symptoms.

More broadly, our results may suggest a different disease process at least in a subset of individuals. At minimum, these distinctions suggest a need to better understand differences and similarities between depression following COVID-19, and typical major depressive disorder.

## Data Availability

Aggregated data are available from the authors on request.

https://covidstates.org

## Acknowledgements

This study was supported by the National Science Foundation (Drs. Ognyanova, Lazar, and Baum). Dr. Perlis is supported by NIMH R56MH115187 and R01MH116270. The sponsors did not contribute to design and conduct of the study; collection, management, analysis, and interpretation of the data; preparation, review, or approval of the manuscript; and decision to submit the manuscript for publication. Dr. Perlis had full access to all the data in the study and takes responsibility for the integrity of the data and the accuracy of the data analysis.

## Disclosures

Dr. Perlis has received consulting fees from Burrage Capital, Genomind, RID Ventures, and Takeda. He holds equity in Outermost Therapeutics and Psy Therapeutics. The other authors report no disclosures.

## Figure and Tables

**SF1.**
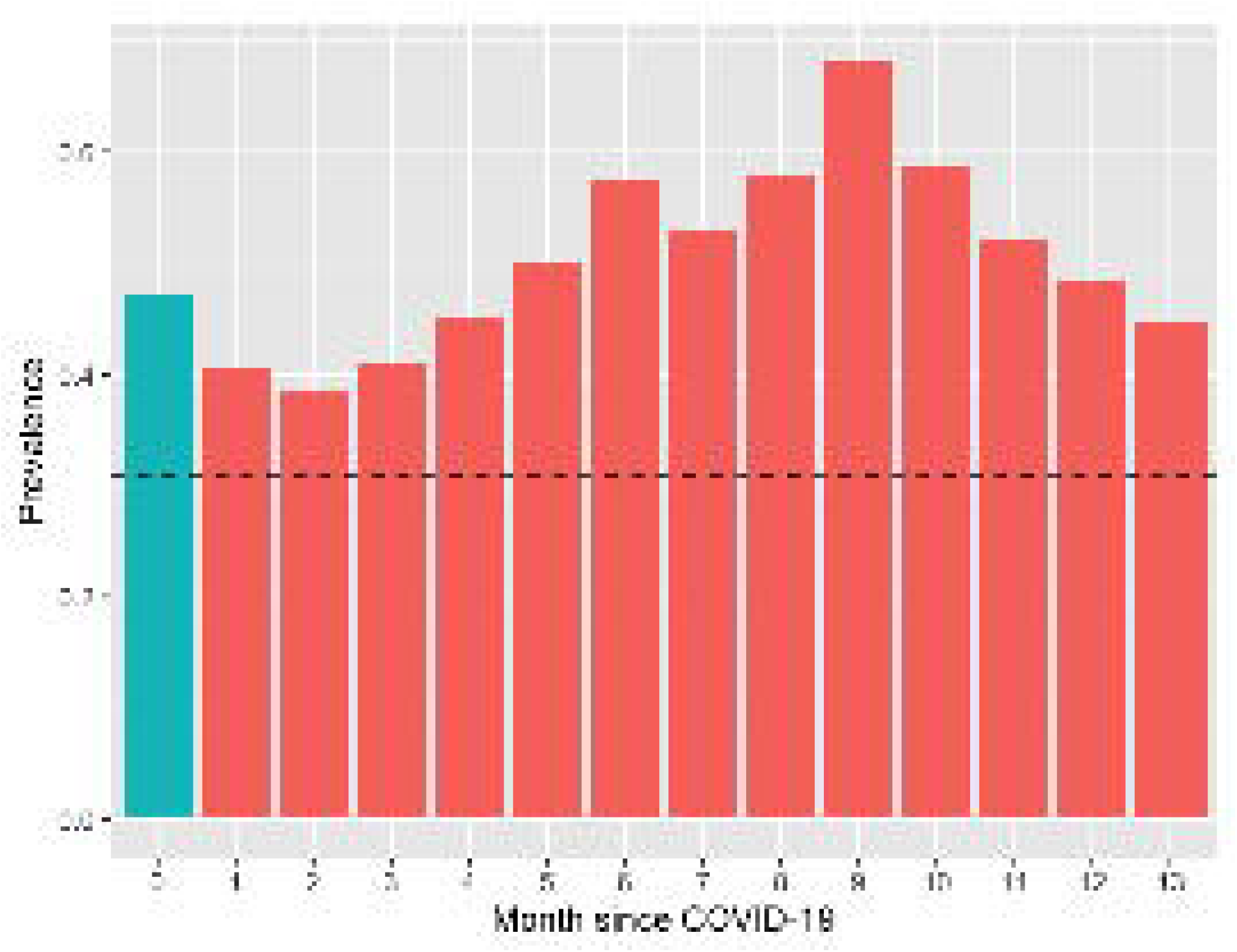
Prevalence of moderate or greater depression, by months since acute COVID-19 illness. For reference, dotted line indicates prevalence of moderate or greater depression among individuals surveyed in May-June 2020.

## Notes

### Author Declarations

Harvard University IRB - exempt

